# The effectiveness of Non Pharmaceutical Interventions in reducing the outcomes of the COVID-19 epidemic in the UK, an observational and modelling study

**DOI:** 10.1101/2020.12.16.20248308

**Authors:** G. Galanis, C. Di Guilmi, D.L. Bennett, G. Baskozos

**Author notes:** Corresponding author: Georgios Baskozos, Nuffield Department of Clinical Neuroscience, John Radcliffe Hospital, University of Oxford, Oxford, UK. contributed equally to this work (G. Galanis, G. Baskozos).

## Abstract

Epidemiological models used to inform government policies aimed to contain the contagion of COVID-19, assume that the reproduction rate is reduced through Non-Pharmaceutical Interventions (NPIs) leading to physical distancing. Available data in the UK show an increase in physical distancing before the NPIs were implemented and a fall soon after implementation. We aimed to estimate the effect of people’s behaviour on the epidemic curve and the effect of NPIs taking into account this behavioural component. We have estimated the effects of confirmed daily cases on physical distancing and we used this insight to design a bevavioural SEIR model (BeSEIR), simulated different scenaria regarding NPIs and compared the results to the standard SEIR. Taking into account behavioural insights improves the description of the contagion dynamics of the epidemic significantly. The BeSEIR predictions regarding the number of infections without NPIs were several orders of magnitude less than the SEIR. However, the BeSEIR prediction showed that early measures would still have an important influence in the reduction of infections. The BeSEIR model shows that even with no intervention the percentage of the cumulative infections within a year will not be enough for the epidemic to resolve due to a herd immunity effect. On the other hand, a standard SEIR model significantly overestimates the effectiveness of measures. Without taking into account the behavioural component the epidemic is predicted to be resolved much sooner than when taking it into account.

## Introduction

The spread of severe acute respiratory syndrome coronavirus 2 (SARS-CoV-2), the virus responsible for COVID-19 has led to more than 9,277,214 confirmed cases and more than 478,691 deaths as of 25/06/020 [1]. Apart from the health-related implications, COVID-19 has been affecting almost every aspect of people’s lives and these effects have been unequally distributed. [2]

Given the current absence of a vaccine, governments have resorted to health policies known as Non-Pharmaceutical Interventions (NPIs), which are aimed to reduce the average number of contacts between individuals (physical distancing). A number of studies have explored the effects of NPIs on the contagion dynamics of COVID-19. [3–6] One of the key focuses of compartmental epidemiological models used is the estimation of the effects of NPIs on the rate of spread of the epidemic, which is captured by the basic reproduction number initially and the effective one later. Hence, the effectiveness of the different policies depends on how the various announced measures reduce this parameter. In order to be able to capture the level of this effect, it is necessary to estimate the value of the effective reproduction number, which in standard compartmental models is assumed to be initially constant and to change as a response to NPIs. [7]

However, data which capture mobility levels of individuals show that in a number of countries, including the UK, people reduced the number of visits and duration of stays (which are related to physical distancing practices) before the NPIs are made and in excess of these measures. This behaviour, fits well with a number of epidemiological models, which take into account behavioural changes over and above NPIs [8–10] hence the effective reproduction becomes (at least partly) endogenous. However, these works are theoretical and have not been applied to data sets related to COVID-19 so far.

The purpose of our study was threefold: (i) assess the influence of NPIs on physical distancing in the UK, taking into account individual behaviour due to observed cases; (ii) extend the standard SEIR model in order to incorporate individual behaviour using the analysis from (i); and (iii) use this extended model to study the effectiveness of the NPIs, including the level and timing of measures taken and the possible effects of lifting the measures.

## Methods

### Data and Statistical Analysis

In order to assess the influence of both NPIs and the observed information, we analysed the correlation between individuals’ mobility levels and the number of daily confirmed cases of the previous day as reported in the WHO dashboard implemented by John Hopkins University, [1] for three different periods: (a) up to the point when advice for physical distancing and avoidance of unnecessary interactions and travel was given (b) between this advice and enforceable lockdown, and (c) after lockdown. Enforceable lockdown includes NPIs ranging from the closure of public spaces, transportation hubs and shops to forbidding interactions with people outside one’s household and ban any unnecessary travel.

Following Buckee et al. (2020), [11] we created an aggregated data time series for physical distancing in the UK using data from Google’s “COVID-19 Community Mobility Reports”, which shows the changes in mobility in six different categories:

1. Retail and recreation, reporting the mobility trends for places such as restaurants, cafés, shopping centres, theme parks, museums, libraries and cinemas.
2. Supermarket and pharmacy, capturing the trends for places such as supermarkets, food warehouses, farmers markets, specialty food shops and pharmacies
3. Parks, which shows the mobility trends for places like national parks, public beaches, marinas, dog parks, plazas and public gardens.
4. Public transport, which shows the mobility trends for places that are public transport hubs, such as underground, bus and train stations.
5. Workplaces, capturing mobility trends to places of work
6. Residential, which shows mobility trends for places of residence.

We noted that not all of the above categories are relevant for measuring levels of physical distancing, which on one hand are related to both NPIs and to individuals’ behaviour, while on the other are relevant for the contagion dynamics. For this reason, we used the categories “Workplaces”, “Public transport”, “Retail and recreation”. We defined as mobility m_t_ mobility at time t, a weighted average of these three mobility categories. In order to calculate the different weights, we first matched these categories with the relevant ones from the national travel survey, 2018 [12] which includes the following travel categories: Business, Education, Escort education, Shopping, Personal Business, Visiting friends at private home, Visiting friends elsewhere, Entertainment / public activity, Sport, Holiday, Day trip, Other.

We matched the National Travel Survey categories “Holiday”, “Day trip”, “Entertainment / public activity”, “Shopping”, “Visiting friends elsewhere (than home)” to the “Retail and Recreation” mobility trend; “Commuting”, “Business” and “Personal Business” to the “Workplaces” mobility trend. Additionally, we hypothesised that the “Public Transport / Transit” mobility trend uniformly influences both the above trends. We then computed the relative weights of the above activities with regards to the total activities and mapped these weights to the three mobility categories. This gave us relative weights of 0.38 for “Retail and Recreation”, 0.29 for “Workplaces” and 0.33 for “Public Transport/Transit”.

We observe that mobility over time resembles a logistic distribution and the same is true for new confirmed cases (*c*_*t*_), figure 1.

**Figure 1:**
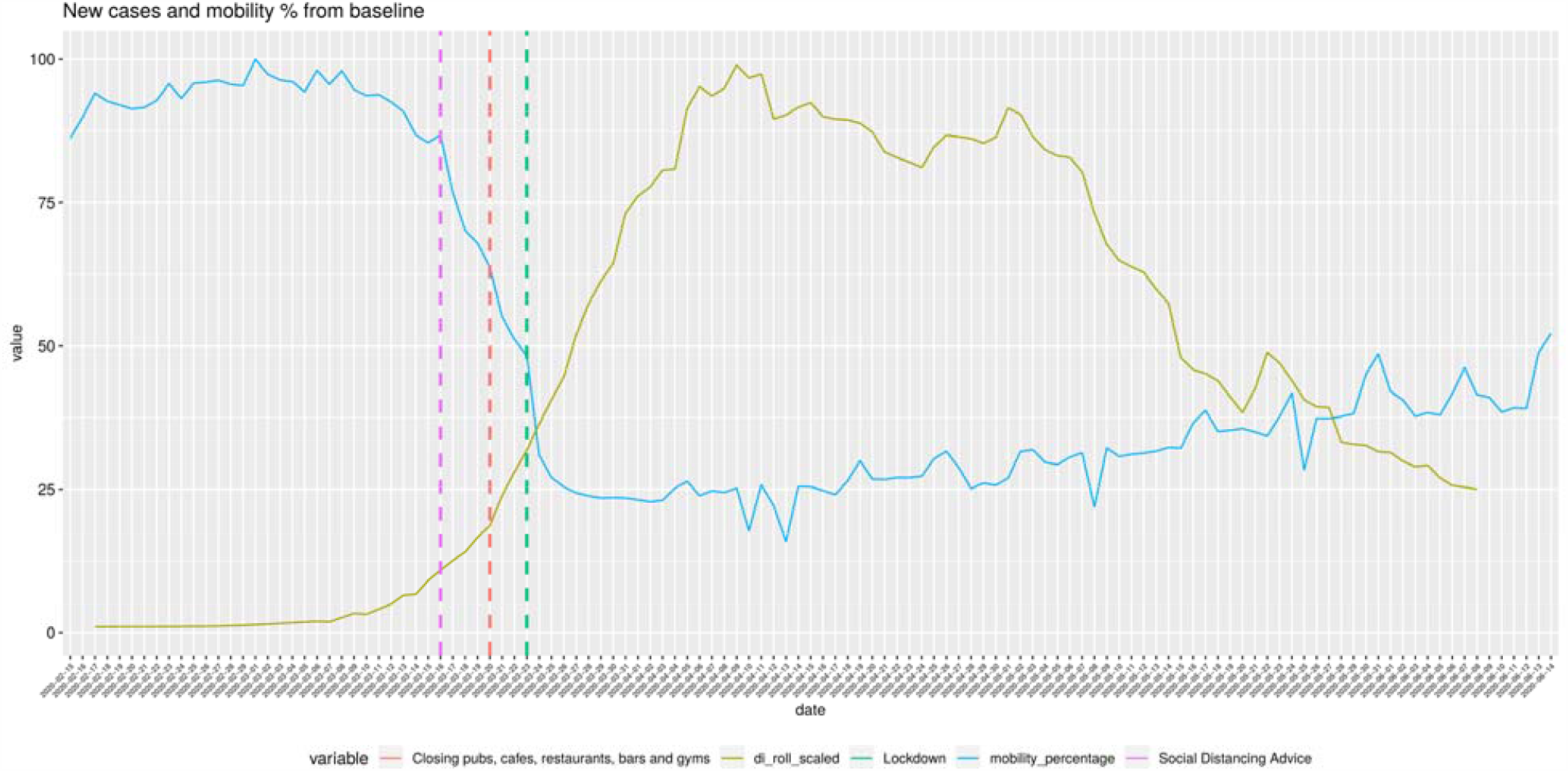
Mobility and new confirmed cases for the UK 15-02-2020: 14-06-2020. The gold line represents the 7-day rolling mean of the new confirmed cases per day. Values are scaled to the range of 0-100. The green line represents the weighted average of the percentage drop of mobility trends from baseline. Vertical dashed lines show the timing of various Non-Pharmaceutical Interventions. Mobility data has been downloaded from google mobility trends and is publicly available, [16] new daily cases has been downloaded form the WHO dashboard [1] and is publicly available.

We observed two periods, one with a sharp decrease in mobility and one with a slight increase. The plots of the confirmed cases follow a very similar pattern but in opposite directions. The different measures taken seem to have an effect of the slope of both lines. The fall in confirmed cases happens around 14 days after the measures have been taken and a big fall in mobility has taken place.

Based on this observation, it is reasonable to assume that there is a linear relationship between the two data series and that the available information in one period affects the decision for the next, which means that m_t_ is a linear function of *c*_*t*-1_ ie.

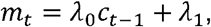

where .*λ* captures the effect of daily confirmed cases and .*λ*_1_ other influences including the effects of NPIs.

However, given that also NPIs affect mobility, we tested this hypothesis for three different periods: before advice, between advice and lockdown, and after lockdown. This hypothesis is supported by very high and significant correlation between the two variables in all three different periods (Supplementary Figure 1).

This observation highlights the fact that measures are not the only factors which influence mobility which in turn is related to physical distancing levels and the reproduction number of COVID-19. Behaviour should therefore be taken into account in relevant models and policy simulations.

### Behavioural SEIR Model

The key variable informing NPIs is the reproduction rate, which is the fraction of the transmission rate of the epidemic (*β*_*t*_) over the recovery rate of infected individuals (*γ*). The daily transmission rate (and hence the basic or effective reproduction rate) directly depends on the number contacts per individual, which means that due to the assumption that mobility is a proxy of daily number of contacts we can express *β*_*t*_ as

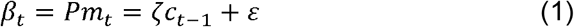

where *P* is the probability of getting infected if contacted by an infected individual, *m*_*t*_ is the mobility as defined above *ε* = *P*.λ_*0*_ and (*ζ*= *P*.λ_1_ capture the relative importance of the behavioural component related to observed infections and the NPIs respectively, such that for *ζ*= 0 only NPIs would affect the level of *β*_*t*_. Note that *ζ* and *ε* are the slope and intercept of the linear model fitting the data as described in the previous section.

We divided the population of N individuals according to the infection status into susceptible (*S*_*t*_), exposed (*E*_*t*_), infected (*I*_*t*_) and removed ones (*R*_*t*_) such that *S*_*t*_ +*E*_*t*_ +*I*_*t*_ +*R*_*t*_ = *N*.

Susceptible subjects might get infected when they contact an infectious individual and if infected, they enter the exposed compartment before the infected one. Following Prem et al. (2020) [13], the infected individuals are split into two further groups where the first group is symptomatic and clinical 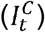 and the second asymptomatic and subclinical 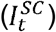. The first group is a fraction *ρ* of the total infected and the second is 1-*ρ* of the total.

Accordingly, the evolution of the infection is given by the following set of dynamic equations

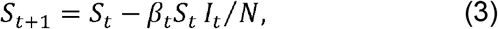

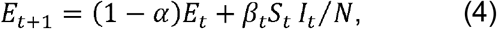

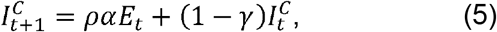

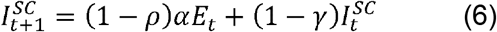

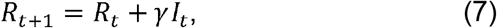

With 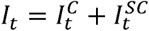.

where *β*_*t*_ captures the daily transmission rate of COVID-19 (as above), *α* is the parameter related to the incubation with 1/*α* being the average incubation period (in days) and y is the daily probability that an infected individual becomes removed (as above). Note that the confirmed daily cases *c*_*t*_ refer to only the symptomatic, which means that *c*_*t*_ = *ρα E*_*t-*1_. hence *equation* (1) can be expressed as

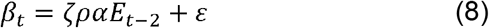

and the model would become a standard (non-behavioural) SEIR model. We note that intuitively *ζ* (should be negative such that an increase in confirmed cases leads to an increase in physical distancing practices, hence to a reduction in *β*_*t*_.

Equations (2)-(8) constitute the baseline Behavioural SEIR based on UK data. The last equation captures the behavioural part of the compartmental model based in the UK and may differ for other countries or other epidemics as the linear relationship which we observed may not hold in other jurisdictions or countries.

## Results

We simulated the contagion dynamics in the UK and the different effects of policies over two periods of 200 and 300 days, respectively, with the demographic changes being ignored, hence keeping the total number of individuals as constant. For equations (3)-(7), we used values from relevant published works. We used the data from a counterfactual SEIR model with *β* = *β*_*0*_, γ = 0.133, ρ = 0.4 (Prem et al. 2020) [13],N = 50000000 (roughly the number of adults in the UK [17]), and started with 1000 people exposed at t=0.

We calibrated the constants of equation (8) as follows:

For *E*_*t-2*_ *= 0*, we get *β*_*0*_ = *ε*. Equation (1) can be expressed as*R*_*0*_ *γ*= *Pm*_*0*_, which means that for *m*_*0*_ = 1 (equivalent to 100% mobility levels) and given values of *λ* and *R*_*0*_ we can find *P* and *β*_*A*_, the value of the reproduction number at the time when the measures are taken.

From Flaxman et al. (2020) [14], we set *R*_*0*_ = 3.8, which gives *β*_*0*_ = *ε* =*R*_*0*_ λ = 0.5. We observed that the UK government started implementing measures when new cases per day were 409 [1]. Using the counterfactual behavioural SEIR model, we chose the time step with the closest value, which was 441 on day 24. Hence, for ρ *αE*_t-2_= 441, *β*_*A*_ = 441 *ζ* + *β*_*0*_ which gives *ζ* = (*β*_*A*_ - *β*_*0*_)/409 = -0.00016

We then considered the peak in new clinical cases per day as the time of the maximum physical distancing and *m*_*t*_ = 0.2 and thus the lowest value of the effective reproduction number. Based on this we calculated the value of *β*_*t*_ on that day (at t=104), which we call *β*_*B*_ and is 0.10. Using this value, we calculated the parameters of equation (8) after the measures. Call these parameters (*ε* ′ = -0.0001734341and E’ = 0.1769653

Using these values, our extended behavioural SEIR model was able to reproduce the dynamics of contagion in the UK (Figure 2 A, B).

**Figure 2:**
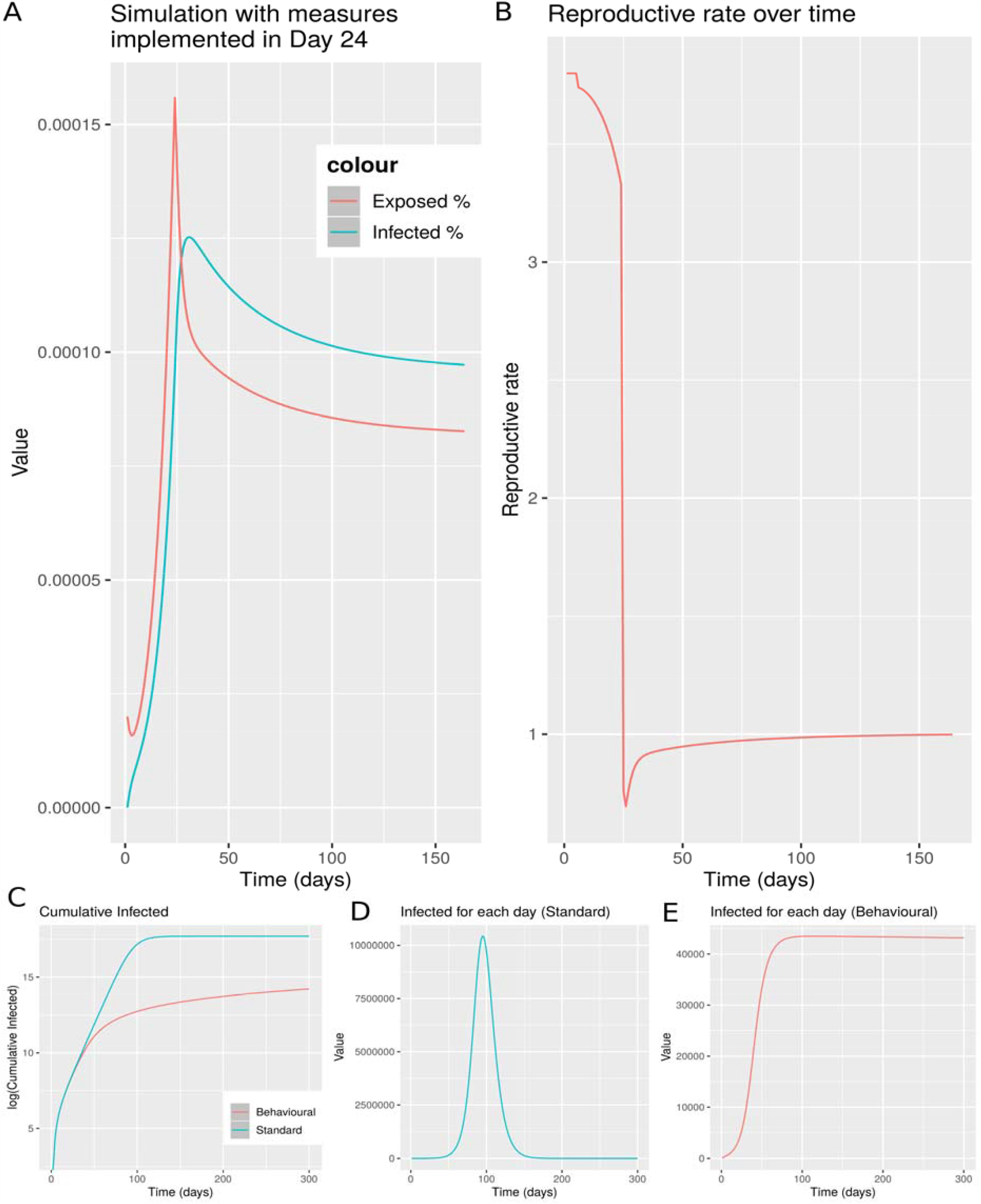
Behavioural SEIR model simulation for the UK adult population, with measures implemented on Day 24, when the number of new observable cases per day is similar to what was in the UK on 16-03-2020. Panel A shows the percentage of Exposed and Infected (both clinical and sub-clinical) individuals per day. Panel B shows the reproductive rate of the disease (Rt) over time. Panel C shows the cumulative infected people as predicted with the standard SEIR model – blue line vs the behavioural SEIR model – red line. Panel D shows the infected people at any time point as predicted by the standard SEIR. Panel E shows the infected people at any time point as predicted by the behavioural SEIR.*R*_*0*_ =3.8, α=0.15625, γ=0.1331221, starting seed = 1000 individuals exposed on Day 0.

We compared the simulation results of our model with a standard SEIR (without the behavioural component) and found that the latter model predicts a number of infections both cumulative (figure 2 C) and per day, much higher than the behavioural one (figure 2 D,E). The difference regarding infections is several orders of magnitude, which highlights the importance of taking into account behavioural factors.

We compared the effectiveness of NPIs in our model and in the standard SEIR. As expected in the standard SEIR the cumulative number of infected individuals is lower than in the behavioural one (figure 3 A). As in the previous case, the cumulative number differs by several orders of magnitude. We not that reproduction rate on the standard SEIR is only affected by NPIs (figure 3 B). Interestingly, we noticed that the maximum number of infected individuals at any point in time using the behavioural SEIR model is significantly and much more realistically lower than the standard SEIR (figure 4 C, D), where it also takes longer for the number of infected to be reduced.

**Figure 3:**
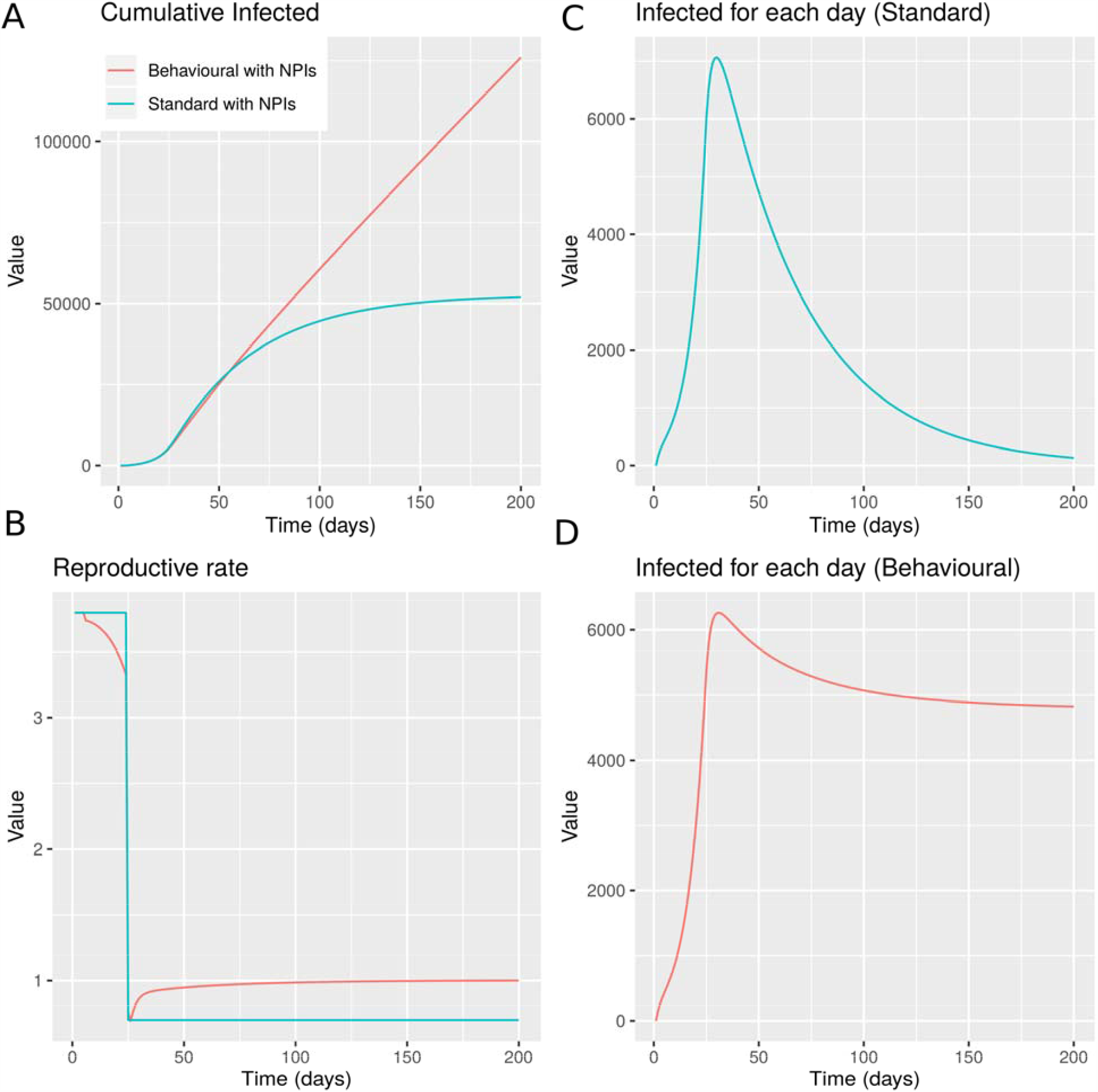
Behavioural SEIR vs standard SEIR with NPIS implemented on Day 24. Panel A shows the cumulative infected people as predicted with the standard SEIR model – blue line vs the behavioural SEIR model – red line. Panel B shows the reproductive rate over time. Panel C shows the infected people at any time point as predicted by the standard SEIR. Panel D shows the infected people at any time point as predicted by the behavioural SEIR.

**Figure 4:**
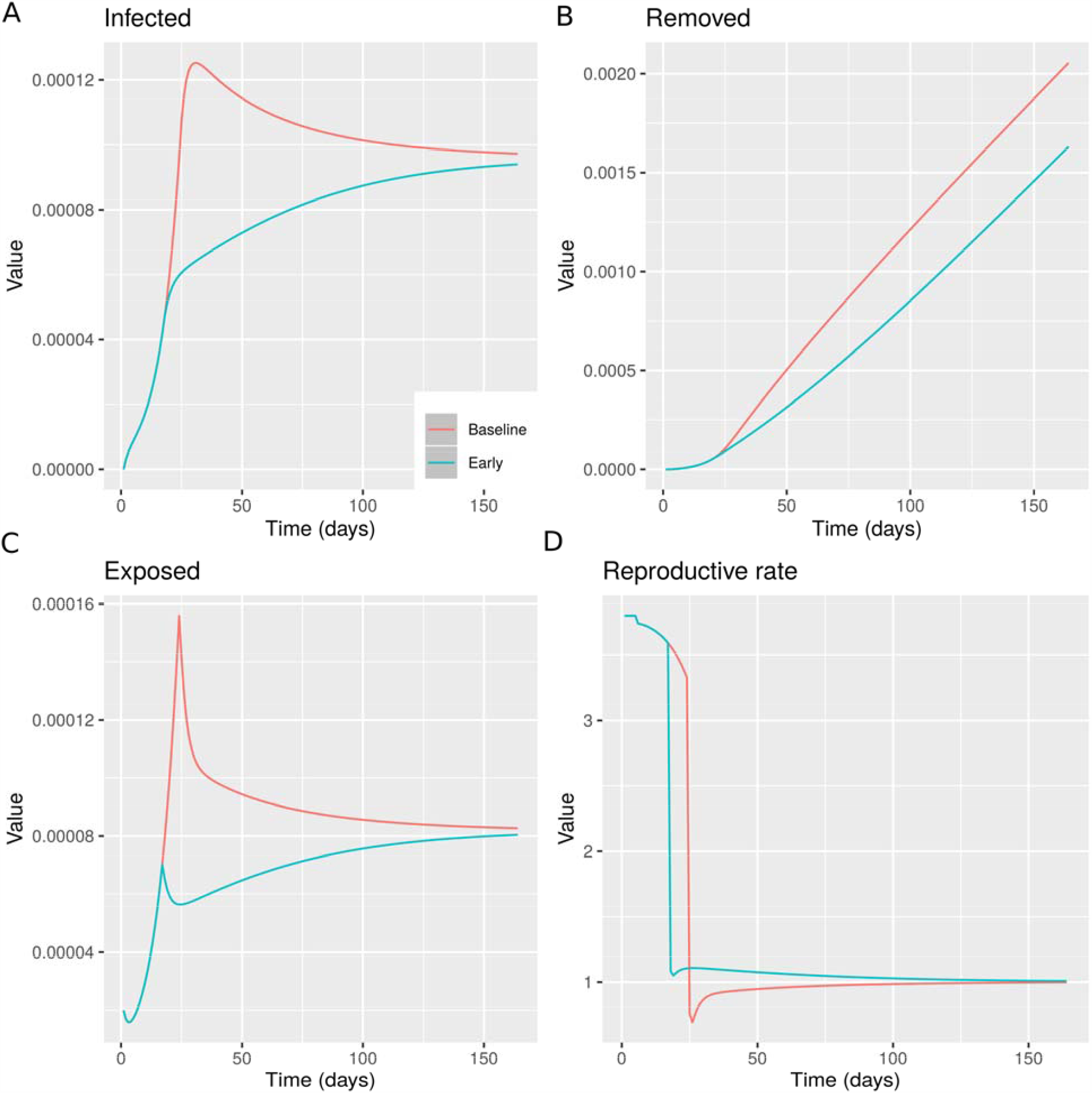
Behavioural SEIR model comparing the effect of measures taken on Day 24, Baseline model – red line or measures taken on Day 17, Early model – blue line. A: Percentage of infected individuals. B: Percentage of Removed individuals. C: Percentage of Exposed individuals. D: Reproductive rate.

We tested the impact on the total number of infected individuals and the maximum confirmed daily cases of the delay of (i) implementing the measures (ii) and lifting restrictions. This allowed us to compute the cost in terms of lockdown days in order to reach the same reproduction number. The next graph shows the dynamics of key variables compared to a hypothetical situation when the same measures had been taken 7 days earlier than the actual date of intervention.

We noticed that, while the timing of measures has an important impact on the number of infected individuals (both daily and cumulatively, figure 4 A, B, C), the reproduction rate is reduced relatively less compared to the scenario where the measures are taken later (figure 4 D). This highlights that all other factors being equal, it may be optimal to have a relatively higher reproduction rate with a lower number of infected rather than the opposite. This is due to the fact that the number of infected individuals at any point in time depends both in the reproduction rate and the number of infected in the previous period. Hence a later intervention would require a higher reduction in the reproduction number to have the same reduction in infections to an earlier one.

We tested the impact of lifting the measures earlier rather than later and also compared this to the hypothetical case of earlier timing of NPIs. As expected, the most efficient policy would be to both delay lifting physical distancing measures and implementing NPIs early (figure 5). We noticed that taking the timing regarding on when the measures are lifted plays a less important role (assuming that there will be a lift) compared to the timing of imposing the measures.

**Figure 5:**
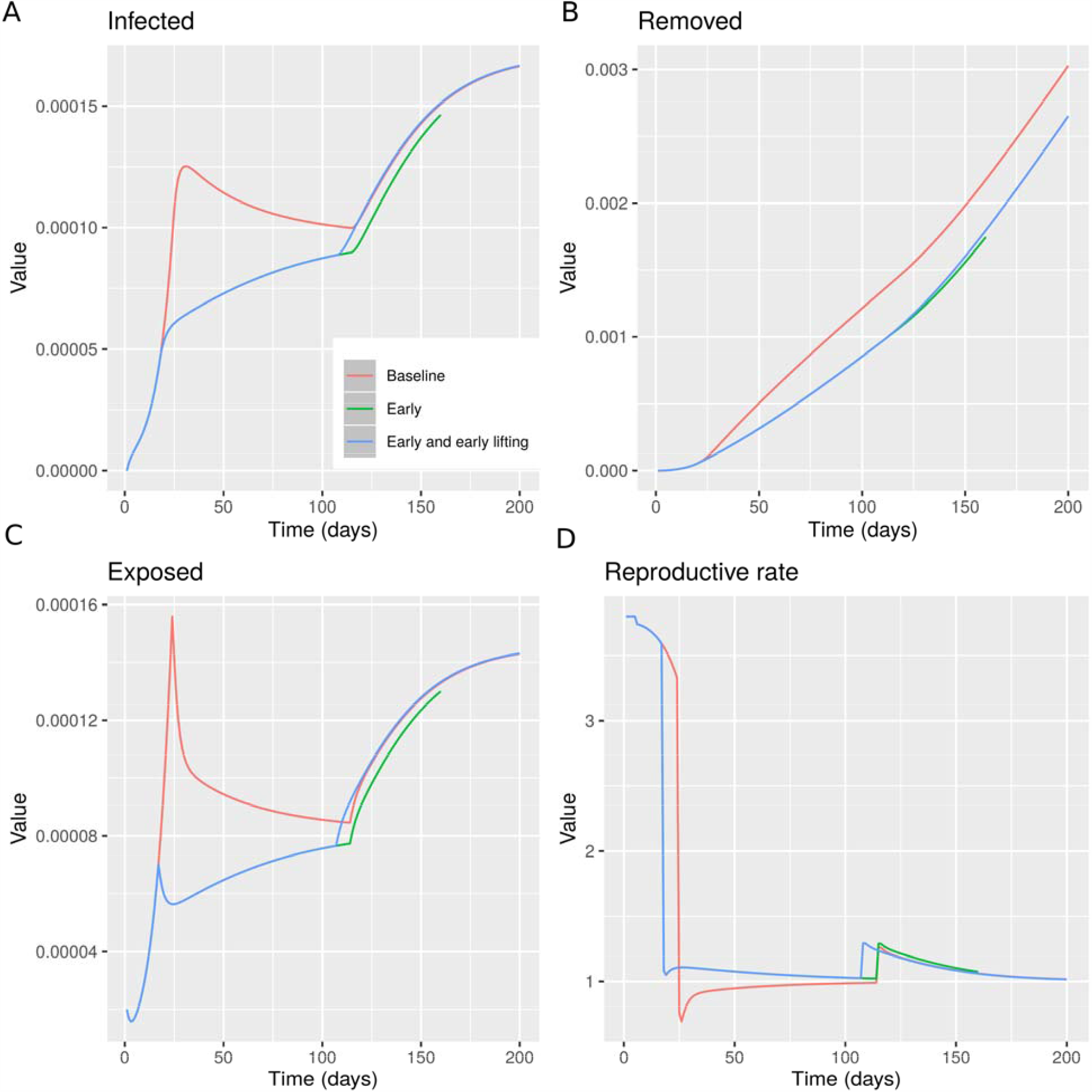
Behavioural SEIR model comparing the effect of measures taken on Day 24 and lifted on Day 114, Baseline model – red line, measures taken on Day 17 and lifted on Day 114, Early model – green line, or measures taken on Day 17 and lifted on Day 107, Early model and early lifting– blue line. A: Percentage of infected individuals. B: Percentage of Removed individuals. C: Percentage of Exposed individuals. D: Reproductive rate.

## Discussion

The absence of a vaccine for SARS-CoV-2 has led most countries to result in NPIs in order to reduce the contagion of COVID-19 and a fast-growing literature has been focusing on estimating the effectiveness of these interventions. We argued that in order to be able to evaluate the interventions, it is necessary to explicitly take account of the behavioural change with regards to physical distancing due to both the relevant NPIs and independent individual choices. If individual behaviour is not taken into account, the levels of physical distancing without measures can be underestimated and similarly the effectiveness of measures can be overestimated. Hence, incorporating an autonomous element of physical distancing can also increase the accuracy of modelling predictions.

Using aggregate mobility data for the UK, we observed that individual mobility levels had been reducing before the measures were taken and have been increasing even before the announcement of relaxation of the measures. We tested whether information regarding confirmed cases can explain the changes in mobility within the different periods of NPIs. In order to also take into consideration, the effects of policies, we considered three distinct periods: before advice, between advice and lockdown, and after lockdown. We found high correlation in all three periods, which confirms the fact that people have been making physical distancing choices using the available information regarding the number of cases which are also assumed to be correlated to the number of deaths. We note that the number of cases reported, at the early phases of the epidemic at least, was a gross underestimate of the real cases. However, the number of deaths has been used in order to infer the number of cases by imposing a somewhat arbitrary death rate. Given this as long as the number of deaths is a fraction of the number of cases the outcome of our behavioural SEIR model will not change and in addition we acknowledge that people make decisions based on imperfect information.

This observation raised two policy related questions with regards to the timing of making the interventions and the time of lifting these. Given that individuals react autonomously, policies are less effective compared to a situation where individuals do not act independently. However, it is not clear how much this behaviour would impact the overall results.

Other studies have taken into account the structured nature of human relationships and the different transmission rates that can be realized in different places, for example care homes or hospitals can show higher transmission rates than households. We acknowledge that our model provides a higher level of abstraction that doesn’t take this into account.

## Conclusion

In order to be able to assess the effects of the different policies, we first extended the standard SEIR model to a behavioural SEIR. This model incorporates the insights stemming from the previous observations; hence it takes into account the fact that mobility is not only determined by NPIs, but also from the observed information with regards to confirmed daily cases. This incorporates a feedback effect between confirmed cases and average number of contacts between individuals, which means that the reproduction rate is (partly) endogenous.

We calibrated the behavioural SEIR model using epidemiological data from previous relevant studies, values for the reproduction number based in the UK, and UK mobility data. We started our simulations with a seed of 1000 people exposed on Day 0. Genetic analysis has showed 1356 transmission lineages of COVID-19 in the UK. [15] We ran simulations that provided a number of insights which are relevant for policy. When the level of daily infections is relatively low, more strict measures are required in order to achieve high levels of physical distancing. This means that the same level of measures may be less effective in reducing the reproduction rate if these are imposed earlier rather than later. However, this does not mean that early measures are less effective in reducing the overall number of infections as NPIs which are imposed even a week earlier can have an important impact in the reduction of infections.

This finding may give an explanation about the initial spread of the epidemic in countries that were hit first. Given the high uncertainty and the much lower volumes of information the endogenous behavioural component we describe couldn’t have a significant effect in these countries. On the other hand, quickly introduced NPIs in the form of enforceable lockdowns were the only way to reduce the spread of the epidemic.

Our results highlight two issues. First, not taking into account the fact that individuals also react themselves over and above NPIs may lead to very misleading projections with regards to the effectiveness of measures. Second, the fact that even though the reproduction number is a relevant variable for policy purposes, it is not necessarily a measure of success of NPIs. A higher reproduction number with less active cases can be preferable to the opposite can be less challenging to the capacity of a given health system. Of course, the basic reproductive rate of the disease as defined by the biological features of the sars-cov-2 virus is still important as it is predictive of the epidemic curve in the absence of NPIs or changes in human behaviour.

## Data Availability

The data used are available from Google and the WHO

https://www.google.com/covid19/mobility?hl=en

https://covid19.who.int/?gclid=CjwKCAjwltH3BRB6EiwAhj0IUD_ZYpmWYI5f1P9OYsYlJKTajlyFvk5aXAwQNMNIoL82XsVMfF4LExoC8e4QAvD_BwE

## Author contributions

Conceptualisation, Methodology, Investigation, Data curation, Visualisation, Writing original draft: G.G., G.B. Writing-Review and editing G.G., D.C, D.L.B, G.B. All authors approve and commented on the final manuscript.

## Data sharing

All data is publicly available from. [1,12,16] No additional data.

## Ethics

No clinical data from human participants has been used for this study.

## Supplementary Material

**Supplemental Figure 1:**
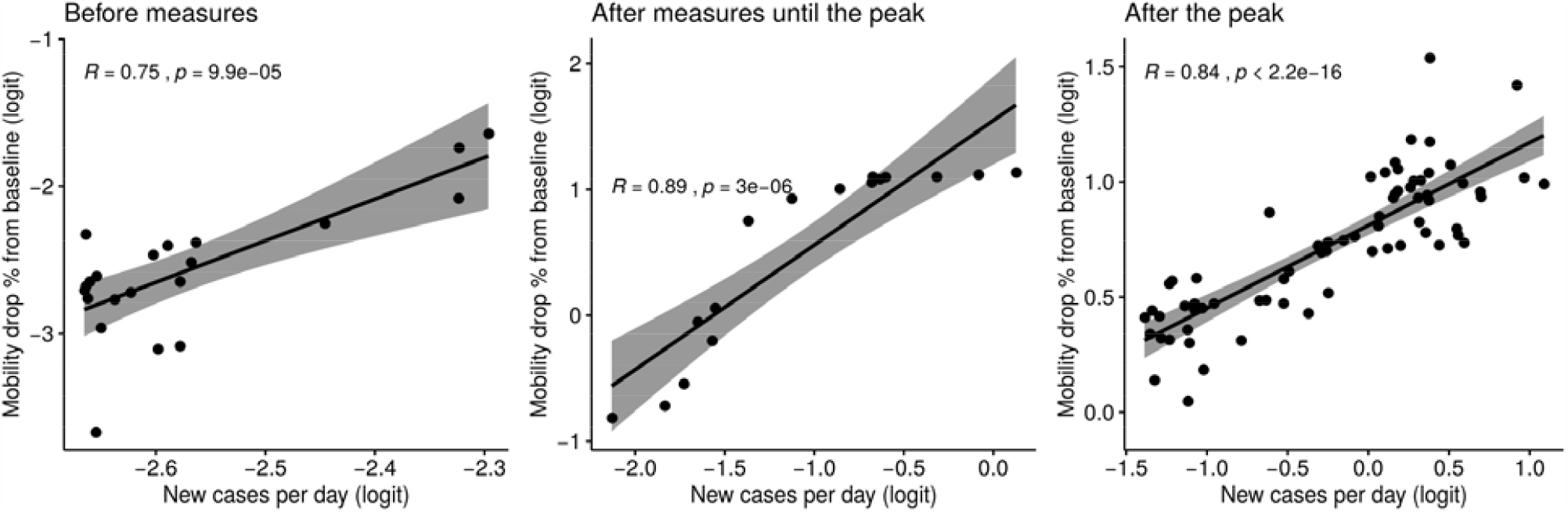
Plots of the logit transformation (ratio of proportions) of the percentage drop of mobility from baseline against new cases per day. Plots show the relationship between mobility and new cases for three periods. A: Before measures, B: After implementing the Non-Pharmaceutical Interventions and C: After the peak of the epidemic. In all cases we have strong significant correlations between drop in mobility and new cases per day. The correlation is stronger after the measures and up to peak of the epidemic.

